# Changes in immunoglobulin levels during clozapine treatment in schizophrenia

**DOI:** 10.1101/2022.05.18.22275238

**Authors:** Kira Griffiths, Maria Ruiz Mellado, Raymond Chung, John Lally, Grant McQueen, Kyra-Verena Sendt, Amy Gillespie, Muhammad Ibrahim, Alex Richter, Adrian Shields, Mark Ponsford, Stephen Jolles, John Hodsoll, Thomas Pollak, Rachel Upthegrove, Alice Egerton, James H MacCabe

## Abstract

**Background:** Clozapine is the only licensed pharmacotherapy for patients with treatment-resistant schizophrenia (TRS), but its use is limited due to adverse effects. Clozapine treatment has been recently associated with reductions in immunoglobulin (Ig) levels cross-sectionally, however prospective studies are required to establish longitudinal effects. This study aimed to determine whether reductions in immunoglobulin levels occur over 6 months after initiating clozapine treatment. An exploratory aim was to investigate relationships between immunoglobulin levels and symptom severity over the course of clozapine treatment.

**Methods:** In 56 participants with TRS, Ig A, M and G levels were measured in serum using a sandwich immunoassay. Samples for analysis were acquired prior to starting clozapine and at 6, 12 and 24 weeks after initiating clozapine treatment. Clinical symptoms were measured using the positive and negative syndrome scale for schizophrenia (PANSS).

**Results:** All three classes of Ig decreased during clozapine treatment. For IgA and IgG the reduction was significant at 24 weeks (IgA: B – 32.7, 95% CI = -61.19, -4.2, p = 0.04; IgG: B – 55.94, 95% CI = -111.03, -0.844, p = 0.05). For IgM the reduction was significant at 12 and 24 weeks (12 weeks: B = -21.73, 95% CI = -37.10, -6.35, p = 0.006; 24 weeks: B = -32.54, 95% CI = -48.89, 16.18, p = 0.0001). Changes in both IgA and IgG were correlated with the percentage change in PANSS total scores over 12 weeks, such that greater reductions in IgA and IgG during clozapine treatment were associated with greater reductions in symptom severity (n = 32, IgA r = 0.59, p = 0.005; IgG r = 0.50, p = 0.02)

**Conclusions:** The observed reductions in immunoglobulin levels over six months of clozapine treatment add further evidence linking clozapine to secondary antibody deficiency. The associations between Ig reduction and symptom improvement may however indicate that immune mechanisms contribute to both desirable and undesirable effects of clozapine.

## INTRODUCTION

Treatment-resistant schizophrenia (TRS) is defined as the persistence of symptoms despite at least two non-clozapine antipsychotic treatment trials of adequate dose, duration and adherence (1). Treatment resistance affects up to 30% of patients with schizophrenia (1), and is associated with worse clinical and functional outcomes (2) and a higher personal, social and economic burden when compared with responsive schizophrenia (3). Clozapine remains the only approved antipsychotic indicated for TRS (4) as it has superior efficacy for this patient group when compared to other antipsychotic medications (5). Whilst the mechanisms underlying improvements in symptoms during clozapine treatment are not fully understood, it has been suggested that immune modulation may play a role in the efficacy of clozapine (6) as well as in its adverse effects. Clozapine is associated with adverse effects involving the immune system, such as severe neutropenia and eosinophilia (7), increased risk of pneumonia (8) and COVID-19 (9). Changes in cytokines (10), neutrophil and platelet count (11) may also occur early during clozapine treatment.

Immunoglobulins (Ig) are an essential component of the adaptative immune system and mediate humoral immunity. Initial observational studies have reported higher rates of clozapine prescription among patients with IgG deficiency (12) and an increased risk of IgM deficiency in patients prescribed clozapine compared to those prescribed non-clozapine antipsychotics (13). A more recent cross-sectional study found that individuals treated with clozapine (n=98) had lower levels of IgA, IgM and IgG compared to those treated with non-clozapine antipsychotics (n=94) (14). Further, that study also reported a significant association between duration of clozapine treatment and reduction in IgG (14). Analysis of a cohort of 1791 individuals assessed for possible antibody deficiency suggested enrichment of clozapine prescription in the 23 participants with a diagnosis of schizophrenia or schizoaffective disorder (15). Clozapine-treatment was also documented in 6 of 7 of individuals with schizophrenia commenced on IgG replacement therapy due to a history of recurrent infection (14). Together, these studies provide circumstantial evidence of an association between clozapine use and risk of antibody deficiency. As reductions in immunoglobulins may increase risk of pneumonia and other adverse effects, these findings could have important implications for monitoring for immunoglobulin deficiency during clozapine treatment. However, prospective studies are first required to establish causal and temporal associations between clozapine use and reductions in immunoglobulins.

The primary aim of this study was to determine whether reductions in immunoglobulins occur over 6 months after initiating clozapine treatment. An exploratory aim was to investigate whether any changes in Ig levels were associated with improvements in symptoms.

## METHODS

### Participants

This study was approved by London South East NHS ethics committee (Ref:13/LO/1857) and all participants provided written informed consent. Patients with TRS who were being initiated on clozapine as part of their clinical care were recruited into the study. Neuroimaging data on a subset of these participants has been reported in previous publications (16) (17), where further details of the study methodology can be found in the online supplementary materials.

Study inclusion required that participants were least 18 years of age and had a clinical diagnosis of schizophrenia (F20) or schizoaffective disorder (F25) according to the International Classification of Diseases Criteria (ICD-10). Inclusion also required that participants were about to commence clozapine initiation as part of their clinical care, and that they were either clozapine naïve or had not taken clozapine in the previous 3 months prior to baseline. Clinical diagnosis was confirmed using the MINI (18). Exclusion criteria included pregnancy, and DSM-IV diagnosis of drug or alcohol dependency, with the exception of nicotine.

### Clinical measures and clozapine plasma levels

Clinical measures were collected using structured clinical interviews at baseline (14 to 0 days before clozapine titration) and repeated at 12 and 24 weeks after clozapine initiation. Symptom severity was assessed using the Positive and Negative Syndrome Scale (PANSS) (19). Clozapine plasma levels were measured in blood samples taken at 6 and 12 weeks and 24 weeks.

### Immunoglobulin analysis

Blood samples for immunoglobulin analysis were acquired at baseline, 6, 12 and 2 -weeks. Samples were centrifuged, aliquoted and stored within the same day at – 80C. Serum levels of IgA, IgM and IgG were determined using the Meso Scale Technology Ig Isotyping Panel 1 Human Kit ®, a sandwich immunoassay.

IgA, IgM and IgG levels were measured simultaneously using a multi-analyte calibrator blend and according to the manufacturer’s instructions. Details of the procedure are available in the Supplementary Methods. Samples were diluted 2×10^−5^ fold and distributed across 96 well plates. A 7-point calibration curve with 4-fold dilution steps plus a zero-calibrator blank was applied to every plate. Calculated concentration was estimated using the raw signal and the calibration curve and was corrected for sample dilution. Each measurement was done in duplicate and the mean of both results was used for analysis.

### Statistical analysis

Statistical analysis was performed using STATA version 16. Summary statistics were used to describe demographic and clinical characteristics of the sample.

The main analysis used linear mixed models with maximum likelihood estimation to examine the effect of clozapine treatment on immunoglobulin levels over time. Linear mixed effect modelling was chosen as it is robust in the presence of unbalanced designs (e.g. missing observations at follow-up due to participant attrition). Three separate models were applied, with level of IgA, IgG or IgM as the primary outcomes of interest. In each model, time was modelled as a fixed effect and treated as a categorical variable (baseline, week 6, week 12, and week 24). A random intercept was included across participants. Further information about the selection of this model is presented in supplementary Table 1. Model residuals were checked via Q-Q plots to assess model assumptions and goodness of fit. Statistical significance was defined as two-tailed p < 0.05.

In an exploratory analysis, we used Pearson’s correlations to investigate whether changes in Ig were associated with percentage change in PANSS total scores after 12 weeks of clozapine treatment. This time period was chosen as it has been associated with clozapine response in approximately 50% of patients (20) (21), and as more data were available for analysis than at the 24-week timepoint. Percentage change in PANSS total score was calculated using the formula: (12-week score – baseline score) / baseline score × 100), after subtracting the minimum possible scores (22).

## RESULTS

Table 1 describes demographic and clinical characteristics of participants. 80% of participants were clozapine naïve prior to baseline. 56 participants had baseline data available for immunoglobulin analysis. Of these, 39 had Ig measures at 6 weeks, 32 at 12 weeks, and 27 at 24 weeks.

**Table 1.**
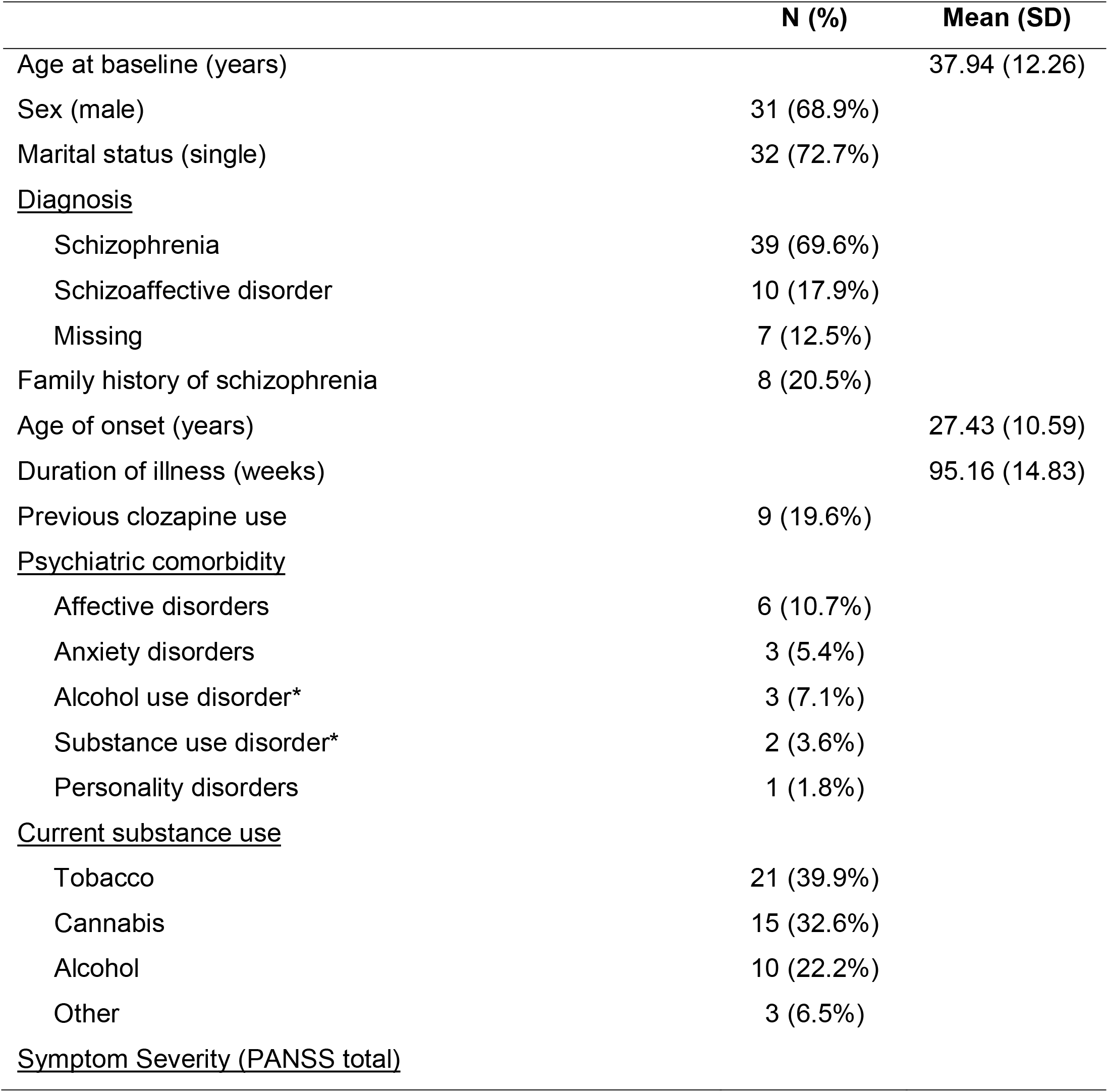
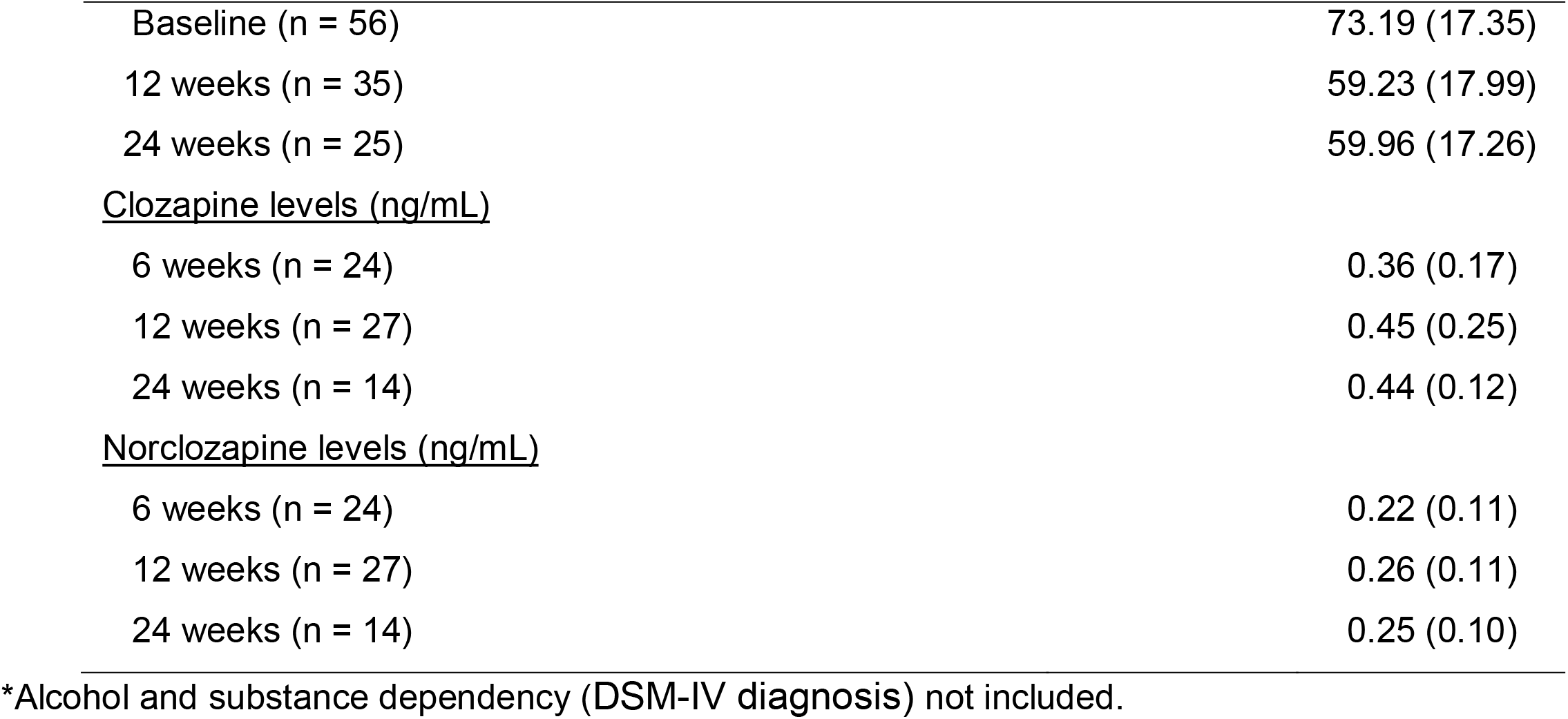
Demographic and clinical characteristics of participants (n=56).

|  | **N (%)** | **Mean (SD)** |
| --- | --- | --- |
| Age at baseline (years) |  | 37.94 (12.26) |
| Sex (male) | 31 (68.9%) |  |
| Marital status (single) | 32 (72.7%) |  |
| <u>Diagnosis</u> |  |  |
| Schizophrenia | 39 (69.6%) |  |
| Schizoaffective disorder | 10 (17.9%) |  |
| Missing | 7 (12.5%) |  |
| Family history of schizophrenia | 8 (20.5%) |  |
| Age of onset (years) |  | 27.43 (10.59) |
| Duration of illness (weeks) |  | 95.16 (14.83) |
| Previous clozapine use | 9 (19.6%) |  |
| <u>Psychiatric comorbidity</u> |  |  |
| Affective disorders | 6 (10.7%) |  |
| Anxiety disorders | 3 (5.4%) |  |
| Alcohol use disorder* | 3 (7.1%) |  |
| Substance use disorder* | 2 (3.6%) |  |
| Personality disorders | 1 (1.8%) |  |
| <u>Current substance use</u> |  |  |
| Tobacco | 21 (39.9%) |  |
| Cannabis | 15 (32.6%) |  |
| Alcohol | 10 (22.2%) |  |
| Other | 3 (6.5%) |  |
| <u>Symptom Severity (PANSS total)</u> |  |  |

|  |  |
| --- | --- |
| Baseline (n = 56) | 73.19 (17.35) |
| 12 weeks (n = 35) | 59.23 (17.99) |
| 24 weeks (n = 25) | 59.96 (17.26) |
| <u>Clozapine levels (ng/mL)</u> |  |
| 6 weeks (n = 24) | 0.36 (0.17) |
| 12 weeks (n = 27) | 0.45 (0.25) |
| 24 weeks (n = 14) | 0.44 (0.12) |
| <u>Norclozapine levels (ng/mL)</u> |  |
| 6 weeks (n = 24) | 0.22 (0.11) |
| 12 weeks (n = 27) | 0.26 (0.11) |
| 24 weeks (n = 14) | 0.25 (0.10) |

\*Alcohol and substance dependency (DSM-IV diagnosis) not included.

### Immunoglobulin levels during clozapine treatment

IgM was significantly decreased at 12 and 24 weeks compared to baseline (12 weeks: β = -21.73, 95% CI = -37.10, -6.35, p = 0.006; 24 weeks: β = -32.54, 95% CI = -48.89, 16.18, p = 0.0001; Table 3; Figure 1). IgA and IgG were significantly decreased at 24 weeks compared to baseline (IgA 24 weeks: β = -32.70, 95% CI = -61.19, -4.21, p = 0.04; IgG 24 weeks: β = -55.94, 95% CI = -111, -0.84, p = 0.05 Table 3, Figure 1). Pearson’s correlation found no significant relationships between Ig levels and clozapine or norclozapine plasma levels at 12 (n = 27) or 24 (n = 14) weeks. There were no significant differences in baseline Ig levels between participants who dropped out of the study compared to those who continued (supplementary Table 2).

**Figure 1.**
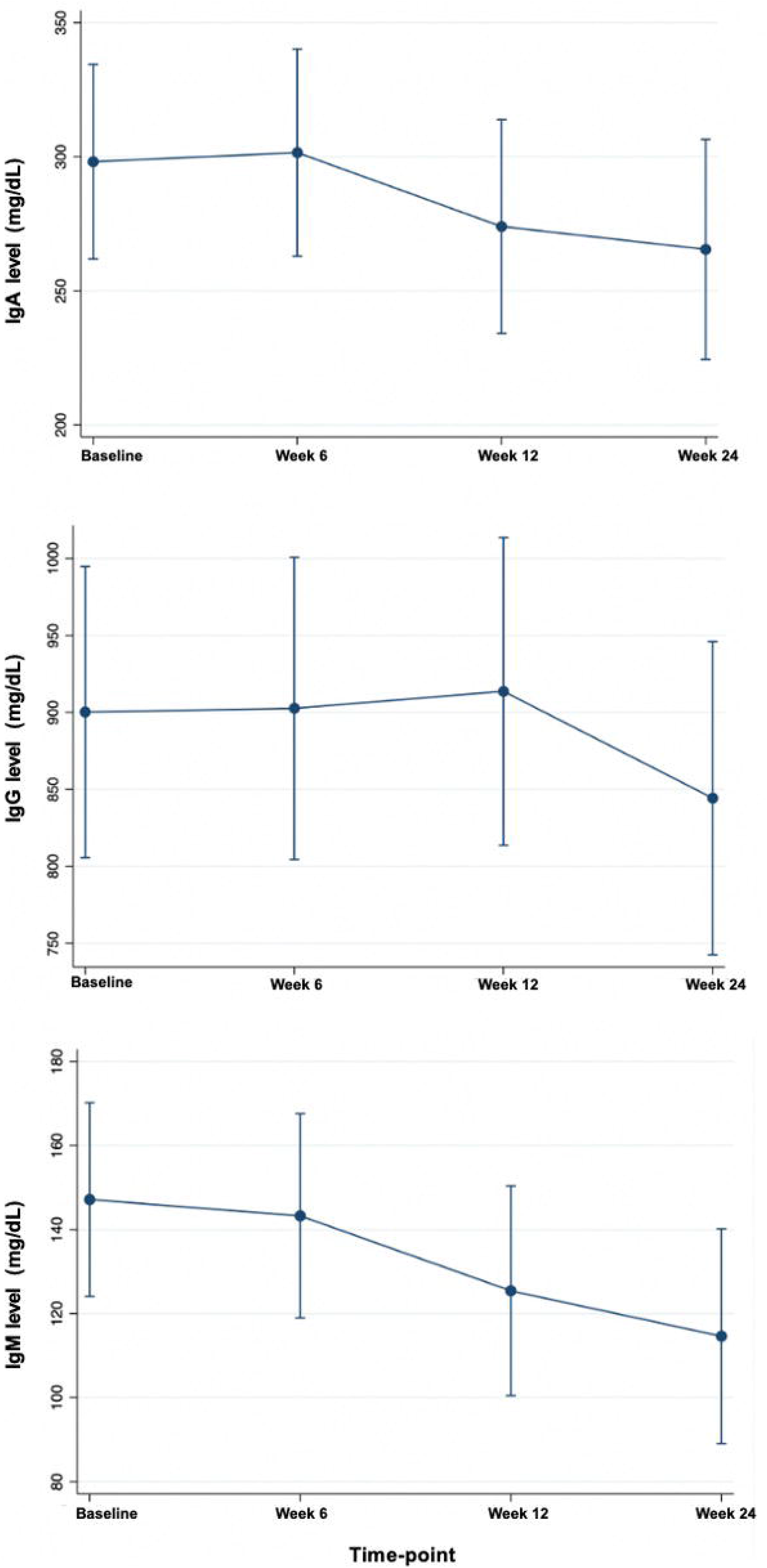
Immunoglobulin levels, expressed as mean ± 95% confidence intervals, over time during clozapine treatment.

As inspection of Q-Q plots for each model indicated a non-normal distribution of residuals, the models were re-run using the logarithmic transformation of immunoglobulin concentrations. The effect of time on IgM at 12 and 24 weeks and the effect of time on IgA at 24 weeks remained significant (IgM 12 weeks: β = -0.18, 95% CI = -0.29, -0.07, p = 0.001; IgM 24 weeks: β = -0.25, 95% CI = -0.37, -0.14, p = < 0.0001 IgA 24 weeks: β = -0.11, 95% CI = -0.20, -0.03, p = 0.009. The effect of time on IgG at 24 weeks became non-significant (β = - 0.048, 95% CI = -0.1, 0.01,p = 0.098).

### Associations between Ig levels and symptoms

Of the 56 participants with immunoglobulin data at baseline, 35 completed structured clinical interviews to assess symptom severity at 12 weeks (Table 1). The correlations between change in IgA and IgG and percentage change in PANSS total scores over 12 weeks were significant, such that greater reductions in IgA and IgG were associated with greater reductions in symptoms (n = 32, IgA r = 0.59, p = 0.005; IgG r = 0.50, p = 0.02).The relationship between change in IgM and percentage change in PANSS total scores was not significant (n = 32, r = 0.12, p = 0.61).

**Table 1.** Estimated marginal means for immunoglobulin levels over time during clozapine treatment

| Immunoglobulin (mg/dL) | Baseline | 6 weeks (n = 39) | 12 weeks (n = 32) | 24 weeks (n = 27) |
| --- | --- | --- | --- | --- |
| **IgA (mg/dL)** | 298.15 [261.88, 334.41] | 301.55 [262.91, 340.18] | 273.97 [234.11, 313.84] | 265.44 [224.41, 306.48] |
| **IgM (mg/dL)** | 147.16 [124.10, 170.22] | 143.27 [118.97, 167.58] | 125.43 [100.48, 150.38] | 114.62 [89.06, 140.19] |
| **IgG (mg/dL)** | 900.22 [805.59, 994.85] | 902.64 [804.47, 1000.80] | 913.64 [813.65, 1013.63] | 844.28 [742.55, 946.02] |

Results are reported as mean [95% confidence intervals]. IgA, immunoglobulin A; IgM, immunoglobulin M; IgG, immunoglobulin G.

## DISCUSSION

To our knowledge, this is the first longitudinal study to measure immunoglobulin levels in patients with treatment-resistant schizophrenia following initiation of clozapine. We found that levels of immunoglobulin classes A, M and G were significantly reduced after 24 weeks of clozapine treatment compared to baseline. These findings extend those from previous cross-sectional studies demonstrating reduced immunoglobulin levels in patients being treated with clozapine compared to non-clozapine antipsychotics (13). Further, we found that the reduction in immunoglobulin G and A levels were positively correlated with reductions in symptom severity. Overall, these results provide initial prospective evidence of a reduction in immunoglobulin levels over the initial early period clozapine treatment.

### Implications

Antibody deficiency is associated with recurrent infections (23). In particular, evidence has shown that clozapine users are at increased risk of pneumonia (24) and antibiotic use (25) when compared to patients who are prescribed non-clozapine antipsychotics. In part, this association might be explained by factors such as sedation (26), sialorrhea (27) and increased rates of smoking (28). Results from the current study provide evidence in support of a mechanism which may reflect impaired humoral immunity, which may drive associations between antibody deficiency and recurrent infections.

Several side effects of clozapine are related to immune system functioning and are closely monitored in clinical practice. For instance, neutropenia is a life-threatening side effect and monitoring programs to detect it promptly have been developed internationally (7). The magnitude of the antibody reduction associated with clozapine has been compared to the reduction observed in users of other immunosuppressive drugs, such as rituximab and methotrexate (25). However, clozapine monitoring programs do not include the routine assessment of immunoglobulins. Early recognition of immunoglobulin deficits could promote timely treatment, which may aid in the prevention of irreversible organ damage, such as bronchiectasis (23). Thus, clozapine users should be considered at increased risk of infection and routine monitoring of immunoglobulin levels and vaccine response in clozapine-treated patients should be considered (25). Regular assessment of immunoglobulin levels is feasible because patients treated with clozapine are routinely monitored as part of clinical practise. Further work on the costs and benefits of such monitoring is warranted.

The finding that a reduction in immunoglobulin levels were associated with a reduction symptom severity is intriguing. Both change in IgA and IgG were associated with change in PANSS total scores over 12 weeks of treatment, such that greater reductions in IgA and IgG were associated with greater reductions in symptom severity. Clozapine is uniquely effective in the treatment of patients with treatment-resistant schizophrenia (4), and there is ample evidence of immune dysfunction in schizophrenia (29). These results raise the possibility that the action of clozapine on the immune system may contribute to its clinical effects and therefore warrant further investigation.

The main finding was a longitudinal reduction in immunoglobulin classes A, M G over the course of clozapine treatment. It is not possible to identify the biological mechanism(s) driving this relationship with the current data. However, possible explanations may include a modulatory effect of clozapine on B or T cell functioning, which in turn impacts immunoglobulin levels. Immunoglobulin production is a complex process that includes recognition of antigens, activation and proliferation of B lymphocytes, differentiation into plasma cells and memory B cells and the secretion of the different classes of antibodies (30). Compared to healthy controls, patients treated with clozapine have more naïve B cell populations, and a reduction in plasmablasts and class-switched B cells (31) (15). There is also evidence that the B-cell profile in some patients treated with clozapine is comparable to patients with common variable immunodeficiency (15); a heterogeneous group of rare diseases associated with intrinsic B cell defects, increased susceptibility to infections, autoimmune manifestations and granulomatous disease (32). In terms of clozapine treatment and T cell functioning, there is some evidence of heightened expression of the DRD3 receptor on peripheral CD4 T cells in clozapine treated patients, as well as reduced counts of regulatory T cells (31). Dopamine antagonism might affect the co-stimulatory loop that facilitates B and T cell interactions (25), however, the impact of clozapine antagonism on peripheral dopamine receptors and whether this is specific or generalisable to other dopaminergic antagonists remains unclear. Going forwards, a prospective study examining the impact of clozapine treatment on B and T lymphocytes changes over time may help to elucidate the underlying biological mechanisms underlying this association between clozapine treatment and reduced immunoglobulin levels.

### Strengths and limitations

A main strength of this study is the prospective design, with the assessment of immunoglobulin levels before and after initiation of clozapine treatment measured within the same participant. This allows longitudinal investigation of the relationship between clozapine use and immunoglobulin levels over time. However, the lack of a non-clozapine comparator group limits causal inference.

Limitations of the study include the lack of a control group, a small sample size, a relatively short observation period and therefore reduced statistical power. Follow-up was incomplete and less than half of the participants had Ig measures at 24 weeks. A slightly higher proportion of participants (57%) had follow-up data at 12 weeks, which reflects the minimum therapeutic time-period suggested to assess clozapine response. As we did not measure infection, the current results do not evidence a direct link between reduction in immunoglobulin levels over clozapine treatment to increased risk of infection. A further limitation is that patients who are prescribed clozapine are frequently treated with adjunctive pharmacotherapies, including other antipsychotics and mood stabilizers, but we were unable to control for these in our analyses.

## Conclusion

This study provides initial prospective evidence of clozapine treatment and reduction in immunoglobulin levels in TRS. This may suggest that clozapine treatment has potential to cause hypogammaglobulinemia. Further studies are required to elucidate the biological mechanisms driving this association.

## Data Availability

All data produced in the present study are available upon reasonable request to the authors.

## REFERENCES

1. Howes OD, McCutcheon R, Agid O et al. Treatment-Resistant Schizophrenia: Treatment Response and Resistance in Psychosis (TRRIP) Working Group Consensus Guidelines on Diagnosis and Terminology. Am J Psychiatry. 2017;174:216–229.

2. Iasevoli F, Giordano S, Balletta R et al. Treatment resistant schizophrenia is associated with the worst community functioning among severely-ill highly-disabling psychiatric conditions and is the most relevant predictor of poorer achievements in functional milestones. Prog Neuropsychopharmacol Biol Psychiatry. 2016;65:34–48.

3. Kennedy JL, Altar CA, Taylor DL, Degtiar I, Hornberger JC. The social and economic burden of treatment-resistant schizophrenia: a systematic literature review. Int Clin Psychopharmacol. 2014;29:63–76.

4. Kane J, Honigfeld G, Singer J, Meltzer H. Clozapine for the treatment-resistant schizophrenic. A double-blind comparison with chlorpromazine. Arch Gen Psychiatry. 1988;45:789–796.

5. Siskind D, McCartney L, Goldschlager R, Kisely S. Clozapine v. first- and second-generation antipsychotics in treatment-refractory schizophrenia: systematic review and meta-analysis. Br J Psychiatry. 2016;209:385–392.

6. Røge R, Møller BK, Andersen CR, Correll CU, Nielsen J. Immunomodulatory effects of clozapine and their clinical implications: what have we learned so far. Schizophr Res. 2012;140:204–213.

7. Remington G, Lee J, Agid O et al. Clozapine’s critical role in treatment resistant schizophrenia: ensuring both safety and use. Expert Opin Drug Saf. 2016;15:1193–1203.

8. Data From the World Health Organization’s Pharmacovigilance Database Supports the Prominent Role of Pneumonia in Mortality Associated With Clozapine Adverse Drug Reactions. [editorial]. Schizophr Bull 2020;46(1):1.

9. Govind R, Fonseca de Freitas D, Pritchard M, Hayes RD, MacCabe JH. Clozapine treatment and risk of COVID-19 infection: retrospective cohort study. Br J Psychiatry. 2021;219:368–374.

10. Kluge M, Schuld A, Schacht A et al. Effects of clozapine and olanzapine on cytokine systems are closely linked to weight gain and drug-induced fever. Psychoneuroendocrinology. 2009;34:118–128.

11. Blackman G, Lisshammar JEL, Zafar R et al. Clozapine Response in Schizophrenia and Hematological Changes. J Clin Psychopharmacol. 2021;41:19–24.

12. Jolles S, Borrell R, Zouwail S et al. Calculated globulin (CG) as a screening test for antibody deficiency. Clin Exp Immunol. 2014;177:671–678.

13. Lozano R, Marin R, Santacruz MJ, Pascual A. Selective Immunoglobulin M Deficiency Among Clozapine-Treated Patients: A Nested Case-Control Study. Prim Care Companion CNS Disord. 2015;17

14. Ponsford M, Castle D, Tahir T et al. Clozapine is associated with secondary antibody deficiency. Br J Psychiatry. 20181-7.

15. Ponsford MJ, Steven R, Bramhall K et al. Clinical and laboratory characteristics of clozapine-treated patients with schizophrenia referred to a national immunodeficiency clinic reveals a B-cell signature resembling common variable immunodeficiency (CVID). J Clin Pathol. 2020;73:587–592.

16. McQueen G, Sendt KV, Gillespie A et al. Changes in Brain Glutamate on Switching to Clozapine in Treatment-Resistant Schizophrenia. Schizophr Bull. 2021;47:662–671.

17. Krajner F, Hadaya L, McQueen G et al. Subcortical volume reduction and cortical thinning 3 months after switching to clozapine in treatment resistant schizophrenia. NPJ Schizophr. 2022;8:13.

18. Sheehan DV, Lecrubier Y, Sheehan KH et al. The Mini-International Neuropsychiatric Interview (M.I.N.I.): the development and validation of a structured diagnostic psychiatric interview for DSM-IV and ICD-10. J Clin Psychiatry. 1998;59 Suppl 20:22-33;quiz 34.

19. Kay SR, Fiszbein A, Opler LA. The positive and negative syndrome scale (PANSS) for schizophrenia. Schizophr Bull. 1987;13:261–276.

20. Rosenheck R, Evans D, Herz L et al. How long to wait for a response to clozapine: a comparison of time course of response to clozapine and conventional antipsychotic medication in refractory schizophrenia. Schizophr Bull. 1999;25:709–719.

21. Meltzer HY, Bastani B, Kwon KY, Ramirez LF, Burnett S, Sharpe J. A prospective study of clozapine in treatment-resistant schizophrenic patients. I. Preliminary report. Psychopharmacology (Berl). 1989;99 Suppl:S68–72.

22. Obermeier M, Mayr A, Schennach-Wolff R, Seemüller F, Möller HJ, Riedel M. Should the PANSS be rescaled. Schizophr Bull. 2010;36:455–460.

23. Patel SY, Carbone J, Jolles S. The Expanding Field of Secondary Antibody Deficiency: Causes, Diagnosis, and Management. Front Immunol. 2019;10:33.

24. Papola D, Ostuzzi G, Gastaldon C et al. Antipsychotic use and risk of life-threatening medical events: umbrella review of observational studies. Acta Psychiatr Scand. 2019;140:227–243.

25. Ponsford MJ, Pecoraro A, Jolles S. Clozapine-associated secondary antibody deficiency. Curr Opin Allergy Clin Immunol. 2019;19:553–562.

26. Clark SR, Warren NS, Kim G et al. Elevated clozapine levels associated with infection: A systematic review. Schizophr Res. 2018;192:50–56.

27. Kaplan J, Schwartz AC, Ward MC. Clozapine-Associated Aspiration Pneumonia: Case Series and Review of the Literature. Psychosomatics. 2018;59:199–203.

28. Caponnetto P, Polosa R, Robson D, Bauld L. Tobacco smoking, related harm and motivation to quit smoking in people with schizophrenia spectrum disorders. Health Psychol Res. 2020;8:9042.

29. Benros ME, Mortensen PB. Role of Infection, Autoimmunity, Atopic Disorders, and the Immune System in Schizophrenia: Evidence from Epidemiological and Genetic Studies. Curr Top Behav Neurosci. 2020;44:141–159.

30. Barahona Afonso AF, João CM. The Production Processes and Biological Effects of Intravenous Immunoglobulin. Biomolecules. 2016;6:15.

31. Fernandez-Egea E, Vértes PE, Flint SM et al. Peripheral Immune Cell Populations Associated with Cognitive Deficits and Negative Symptoms of Treatment-Resistant Schizophrenia. PLoS One. 2016;11:e0155631.

32. Abbott JK, Gelfand EW. Common Variable Immunodeficiency: Diagnosis, Management, and Treatment. Immunol Allergy Clin North Am. 2015;35:637–658.

