## Supplementary Materials for "Changes in immunoglobulin levels during clozapine treatment in schizophrenia"

**SUPPLEMENTARY MATERIAL**

**Supplementary Methods**

Protocol of the Human/NHP Isotyping Kits.

Information was summarised and adapted from Meso Scale Discovery ® Manufacturer’s procedure

1) Sample and reagent preparation.

- Calibrators were put on ice
- 7 calibration solutions were prepared using the manufacturer’s calibrator
  - The stock calibrator was diluted 20-fold in Diluent 100
  - 4-fold dilution steps and zero calibrator preparation
- Samples were diluted 250000-fold using Diluent 100
- Antibody detection solution was prepared by diluting stock detection antibody 50-fold in Diluent 100.
- 2x Read Buffer T was prepared

2) 150 uL per well of Blocker A solution were added. Incubation at room temperature and shaking for 30 minutes
3) Plates were washed 3 times with 150 ug/well of PBS-T. Then 25 uL/wll of diluted sample were added (calibrator or unknown), incubated at room temperature with shaking for 2 hours
4) Plates were washed 3 times with 150 uL/well of PBS-T. 25 uL/well of 1x detection antibody solution was added, followed by 2-hour incubation at room temperature with shaking.
5) Plates were washed 3 times with 150 uL/well of PBS-T. 150 uL of the 2x Read Buffer T were added to each plate and plates were finally read on MSD imager.

**STable 1.** Akaike's Criterion and Bayesian criteria from linear mixed models using fixed effects, random intercept and random intercept and coefficient.

|  | AIC | BIC |
| --- | --- | --- |
| IgA | | |
| Fixed effects | 1961.045 | 1976.23 |
| Random intercept | 1836.437 | 1854.659 |
| Random intercept and random coefficient | 1838.437 | 1859.696 |
| IgM | | |
| Fixed effects | 1832.522 | 1847.707 |
| Random intercept | 1677.425 | 1695.647 |
| Random intercept and random coefficient | 1678.895 | 1700.153 |
| IgG | | |
| Fixed effects | 2230.254 | 2245.439 |
| Random intercept | 2074.28 | 2092.502 |
| Random intercept and random coefficient | 2228.254 | 2240.402 |

Akaike’s information criterion and Bayesian information criterion were used to assess relative goodness of fit of the models. All linear mixed models were fit using maximum likelihood estimation with an unspecified covariance structure. In each model, immunoglobulin level was the dependent variable of interest and time was modelled as a fixed effect. For all three immunoglobulins, the model which included a random intercept for participants had the lowest AIC and BIC.

**Abbreviations:** AIC; Akaike’s information criterion; BIC, Bayesian information criterion; IgA, immunoglobulin A; IgM, immunoglobulin M; IgG, immunoglobulin G.

**STable 2.** Baseline immunoglobulin levels in those who dropped out and those who completed the study

|  | **Drop out (n = 32)** | **Continued (n = 24)** | **p** |
| --- | --- | --- | --- |
| IgA | 291.59 (25.10) | 306.89 (27.98) | 0.69 |
| IgM | 135.26 (81.06) | 163.02 (94.88) | 0.24 |
| IgG | 884.23 (418.90) | 921.54 (306.67) | 0.71 |

Drop out = participants who did not provide a blood sample for immunoglobulin analysis at 8, 12 or 24 weeks. Values are reported as mean (SD). Between group comparisons were performed using ttest.
